# The associations of lifestyle factors with fatigue and the ability to work in the first year after colorectal cancer surgery and rehabilitation

**DOI:** 10.64898/2026.02.17.26346469

**Authors:** T Vlaski, R Caspari, H Fischer, B Bilsing, CM Fernandes-Almeida, M Hoffmeister, M Slavic, K Steindorf, H Brenner, B Schöttker

## Abstract

**Background:** The dynamic associations of lifestyle factors with fatigue and work ability in colorectal cancer (CRC) from pre-diagnosis, over rehabilitation until convalescence in the first year after rehabilitation are largely unexplored.

**Methods:** N = 682 CRC patients were recruited for the MIRANDA cohort study in 4 German rehabilitation clinics. The five-component Healthy Lifestyle Score (HLS; smoking, alcohol, diet, physical activity, BMI) was assessed pre-diagnosis, during rehabilitation (which was up to 12 months after surgery), and 12 months after rehabilitation. Fatigue and the ability to work were assessed during rehabilitation and in 3-month-intervals thereafter.

**Results:** The HLS was rather stable over time, whereas fatigue and ability to work improved in the first 3 months after rehabilitation and remained stable thereafter. Higher HLS points, either assessed prior diagnosis or during rehabilitation, were associated with lower fatigue and better ability to work during in-patient rehabilitation. Compliance with the smoking criterion was the most important factor. Compliance with the physical activity criterion during rehabilitation was also associated with fatigue and ability to work during rehabilitation. In longitudinal analysis adjusted for fatigue and ability to work at rehabilitation, pre-diagnosis adherence to the alcohol consumption criterion was associated with favorable changes of fatigue and ability to work from rehabilitation to 3- and 12-month follow-up. However, the total HLS and other life-style factors were not associated with the outcomes in longitudinal analysis.

**Conclusions:** Addressing lifestyle factors during rehabilitation is an important cornerstone in fatigue management and can improve the ability to work of CRC patients.

## 1. Background

Colorectal cancer (CRC) accounted for approximately two million new diagnoses and close to one million deaths in 2020, and 10.7% of all new cancer cases and 9.5% of cancer deaths were attributed to CRC globally [1]. The survival rates for CRC are increasing, primarily due to earlier detection through enhanced screening protocols and advancements in CRC treatment modalities. Additionally, the demographic shift toward an older population in developed countries is leading to a higher incidence of CRC diagnoses. As a result, the prevalence of CRC survivors is steadily increasing [2].

Cancer-related fatigue is a pervasive and debilitating symptom experienced by many CRC patients, which can persist for months or even years after primary cancer therapy [3]. The cancer and its treatment side effects are the main causes of fatigue [4]. However, fatigue is a multifactorial condition, and a link to inflammation was suggested among its various causes [5]. Cancer-related fatigue is strongly associated with the patient’s quality of life and a reduced ability to work, significantly affecting patients’ professional lives and productivity [6, 7]. Given these implications, it is necessary to investigate interventions that can effectively mitigate fatigue, and thereby improve the quality of life and functional capacity of CRC survivors.

Life-style factors are an area for such interventions. CRC patients engaging in regular physical activity, maintaining a normal BMI, and adhering to diet recommendations have been observed to have lower levels of fatigue [8]. Traditionally, such studies have focused on individual lifestyle factors. This approach fails to capture the complexity of real-life scenarios where lifestyle behaviors are interconnected.

Recognizing this gap, the Healthy Lifestyle Score (HLS) was developed to provide a more comprehensive measure for assessing the combined impact of lifestyle factors on CRC risk and prognosis [9, 10]. It combines five key lifestyle elements (diet, physical activity, alcohol consumption, BMI, and smoking). A study utilizing the HLS found that a change towards a healthier lifestyle is associated with better health-related quality of life in CRC patients [11]. However, fatigue and the ability to work were not used as endpoints in previous studies.

Therefore, the aim of this investigation is to evaluate whether the HLS and its components pre-diagnosis and during rehabilitation are associated with fatigue and the ability to work among CRC patients undergoing rehabilitation. Additionally, we aim to evaluate the longitudinal association between the HLS and its components with changes in fatigue and ability to work after rehabilitation over a follow-up period of up to 12 months.

## 2. Materials and Methods

### 2.1 Data source

The MIRANDA study is a national, multicenter, prospective observational cohort study conducted across Germany. Participants eligible to enroll are CRC patients aged 18 years or older who have completed their primary CRC treatment within the past 12 months and are currently admitted to one of five participating rehabilitation clinics for a minimum of a three-week inpatient rehabilitation program. Every three months during the first year, participants are asked to fill out self-administered follow-up questionnaires. No written consent was the only exclusion criterion [7].

**Figure S1** shows the flow-chart of study participants for this investigation. Participants from this cohort were included if they had completed the questionnaires at recruitment between September 2020 and October 2025 (n=760). Exclusion criteria were: No CRC surgery within 12 months before enrolment, more than one missing lifestyle factor of the HLS, more than 50% items of the FACIT-F-FS missing, or the ability to work information missing (information at rehabilitation). Finally, n=682 participants could be included in the analyses with the outcomes assessed at rehabilitation. In the longitudinal analyses with 3-month follow-up data, n=132 study participants were excluded because they were lost to follow-up, leaving n=550 for analysis. In the analysis using the 12-month follow-up, n=277 study participants could be excluded because n=111 were drop-outs and n=166 were recruited so late that they did not have time to complete the 12-month follow-up questionnaire, leaving n=409 for analysis.

### 2.2 HLS assessment

The HLS comprises of a BMI, smoking, alcohol consumption, diet, and physical activity criterion [9]. Each life-style factor is assigned 0 points or 1 point, and these points are then summed to create an overall score ranging from 0 to 5 points, with 5 points indicating that all 5 lifestyle factors are meeting the criteria for a healthy lifestyle.

In the MIRANDA study, participants’ dietary habits were assessed using a food frequency questionnaire, which evaluated the average frequency of consumption of red meat, processed meat, whole grains, fruits, and vegetables/salads over the previous 12 months. Dietary habits were quantified on a scale from 1 to 50 points. Favourable dietary factors, such as whole grains, fruits, and vegetables were scored positively, while unfavourable factors, such as red and processed meats, were scored negatively. Those achieving a diet score ≥ 34 points were given 1 point in the HLS.

Alcohol intake was estimated based on the reported average weekly consumption of beer, wine, and spirits, taking into account standard German serving sizes and alcohol content of these beverages. Participants were assigned 1 point if their alcohol consumption ≤ 24 g/day for men and ≤ 12 g/day for women [12].

Physical activity levels were quantified in average weekly intensity (light, moderate, vigorous) and frequency (never, three times per week, every day, multiple times a day). Participants who complied with the recommended minimum of at least 75 minutes of vigorous or 150 minutes of moderate physical activity per week received 1 point.

The body mass index (BMI) was calculated from self-reported weight and height. A BMI within the healthy range of >18.5 kg/m^2^ and <25 kg/ m^2^ scored 1 point.

Smoking status was assessed through self-administered questionnaires. Never-smokers or former smokers with a history of less than 30 pack-years were given 1 point.

These criteria are outlined in Table 1.

**Table 1.**
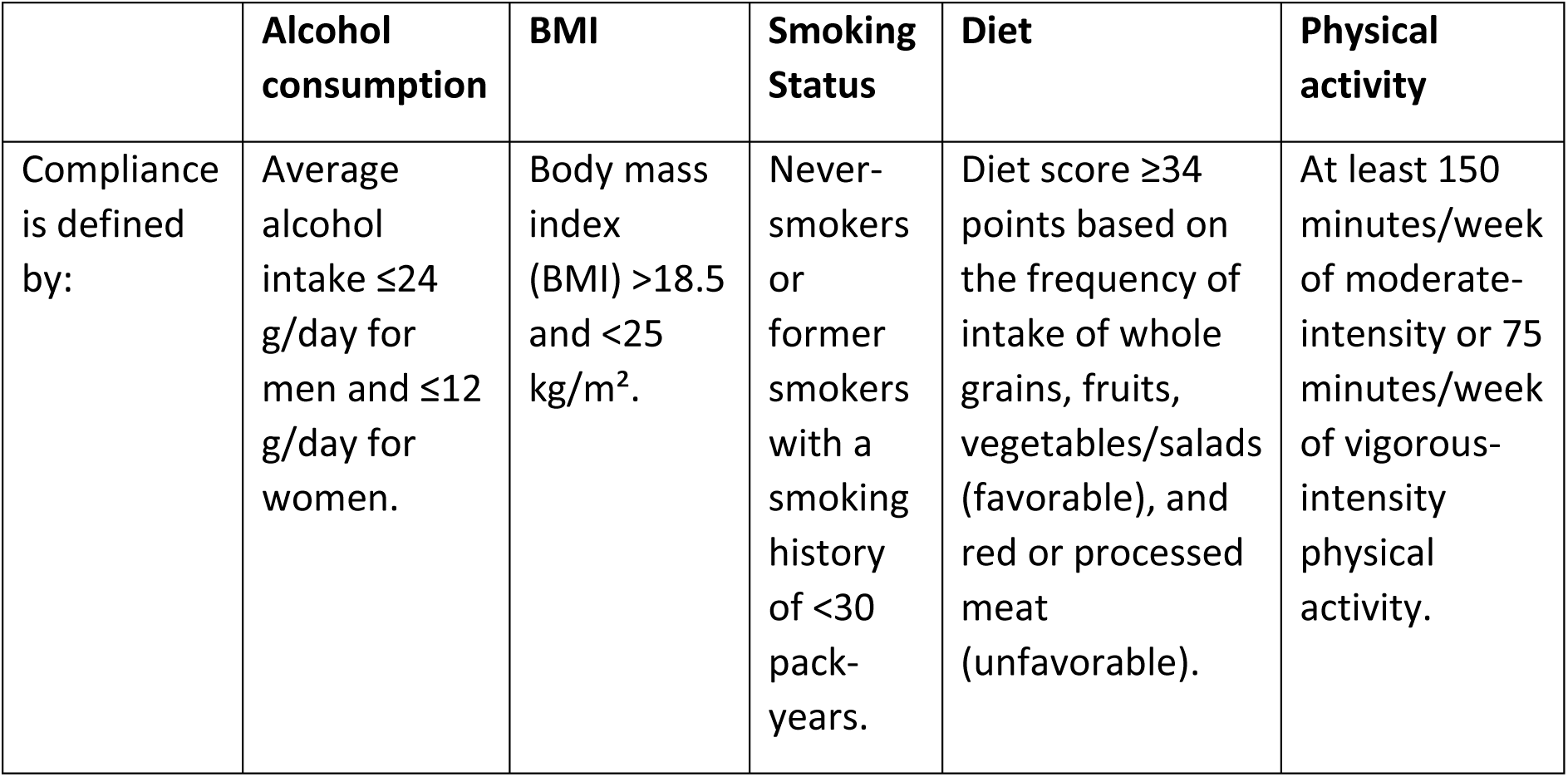
Compliance definitions for each component of the Healthy Lifestyle Score (HLS).

|  | **Alcohol consumption** | **BMI** | **Smoking Status** | **Diet** | **Physical activity** |
| --- | --- | --- | --- | --- | --- |
| Compliance is defined by: | Average alcohol intake $\leq 24$ g/day for men and $\leq 12$ g/day for women. | Body mass index (BMI) $> 18.5$ and $< 25$ kg/m<sup>2</sup>. | Never-smokers or former smokers with a smoking history of $< 30$ pack-years. | Diet score $\geq 34$ points based on the frequency of intake of whole grains, fruits, vegetables/salads (favorable), and red or processed meat (unfavorable). | At least 150 minutes/week of moderate-intensity or 75 minutes/week of vigorous-intensity physical activity. |

The HLS was calculated for three specific time points:

1. Before CRC diagnosis (retrospective questions asked in the self-administered questionnaire during rehabilitation),
2. At rehabilitation, which was up to 12 months after CRC surgery
3. 12 months after rehabilitation

### 2.3 Fatigue and Ability to work

Fatigue was estimated using the Functional Assessment of Chronic Illness Therapy – Fatigue – Fatigue Scale (FACIT-F-FS) score. Cancer-related fatigue was defined as having a score ≤ 34 on the FACIT-F-FS, which has been previously established as clinically relevant fatigue [13].

The ability to work was estimated using the FACIT-F-Functional Well-Being-Ability to Work (FACIT-F-FWB-AW) item. A low-to-moderate ability to work was defined by the response categories “Not at all” and “A little bit” to the FACIT-F-FWB-AW item (“I am able to work, including work at home”) [7]. In a previous analysis from the MIRANDA study, we demonstrated a fair agreement between the FACIT-F-FWB-AW item and the Work Ability Index (WAI) among participants who were working at the 9-month follow-up, supporting the construct validity of the FACIT-F-FWB-AW as a brief measure of work ability in this population [7].

Fatigue and the ability to work were assessed at five time points:

- At rehabilitation, which was up to 12 months after CRC surgery
- 3, 6, 9 and 12 months after rehabilitation

### 2.4 Covariates

Data on sex, age, smoking, physical activity, BMI, pain, number of comorbidities, CRC stage, time since CRC surgery, type of treatment, and number of comorbidities were used as covariates. Data were collected at baseline using a standardized questionnaire.

### 2.5 Statistical Analyses

The trajectories of the HLS from the before CRC diagnosis to 12 months, over rehabilitation to 12 months after rehabilitation were assessed with means and 95% confidence intervals (95%CI). The changes in the distribution of the HLS and its components were additionally shown graphically using bar plots. The changes in mean (95%CI) FACIT-F-FS and FACIT-F-FWB-AW scores in the first year after rehabilitation were also displayed graphically.

Multivariable linear regression analyses were performed to assess the associations of the HLS and its individual components with FACIT-F-FS and FACIT-F-FWB-AW scores. All models were adjusted for age, sex, CRC stage, time since CRC surgery, chemotherapy and/or radiotherapy since CRC surgery, and number of comorbidities as reported at baseline. The longitudinal analyses on fatigue and ability to work changes were additionally adjusted for the initial score distributions in the respective analyses. Four distinct analyses were conducted:

1. Retrospectively assessed pre-diagnosis life-style factors and the fatigue and ability to work at rehabilitation
2. Life-style factors at rehabilitation and the fatigue and ability to work at rehabilitation (cross-sectional analysis
3. Retrospectively assessed pre-diagnosis life-style factors and changes in fatigue and ability to work from rehabilitation to follow-up at 3 months (longitudinal analysis)
4. Retrospectively assessed pre-diagnosis life-style factors and changes in fatigue and ability to work from rehabilitation to follow-up at 12 months (longitudinal analysis)

To impute up to one lifestyle factor of the HLS (for 4.4% of the study population) and missing covariate values, multiple imputation by chained equations (MICE) was applied. Covariate data had low numbers of missing values, with no variable exceeding 1% missingness. Twenty-five data sets were imputed separately with the SAS procedure PROC MI. To our knowledge, missing values were missing at random. All analyses were performed on the twenty-five imputed data sets, and the results were combined using the SAS procedure PROC MIANALYZE. All analyses were done using SAS software version 9.4. A two-sided significance level of p < 0.05 was used for all the tests.

## 5. Results

### 3.1 Description of the study population

Characteristics of the 682 study participants at recruitment during rehabilitation are summarized in **Table 2**. The mean age was 63 years (SD 10.3), and 44.3 % were female. The majority of participants had 10–12 years of school education (42.8%), while 36.7% had ≤9 years and 20.5% ≥13 years of education. CRC stages I, II, and III were represented in similar proportions (35.2%, 32.0%, and 27.5%, respectively), and 5.3% of participants had stage IV cancer. The mean time since CRC diagnosis was 6.8 months (SD 6.7). All participants had undergone surgery before enrolment, with 31.8% having had surgery within the first month prior to baseline assessment. Adjuvant chemotherapy was administered to 43.5% and radiotherapy to 18.3% of participants.

**Table 2.**
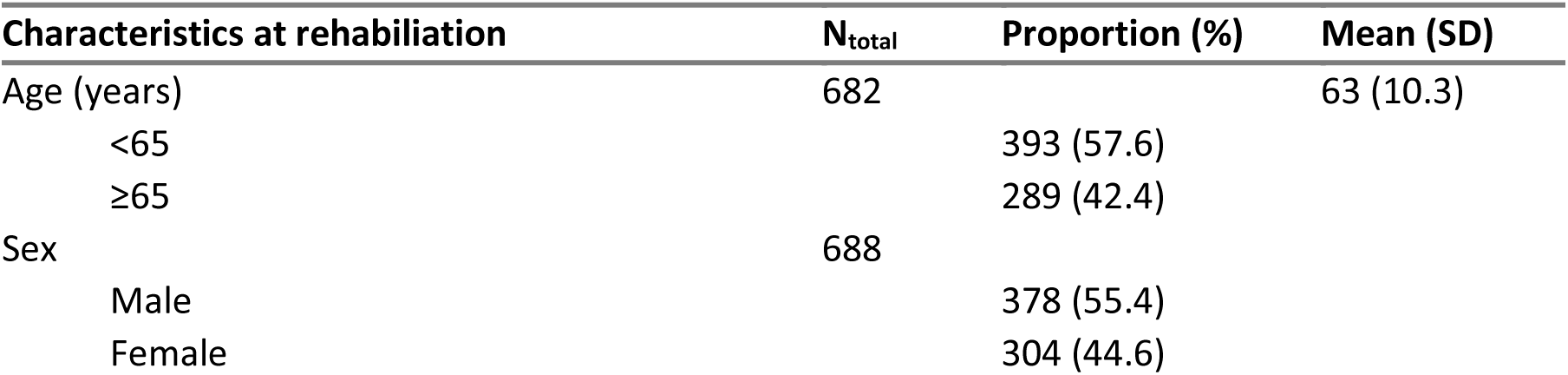
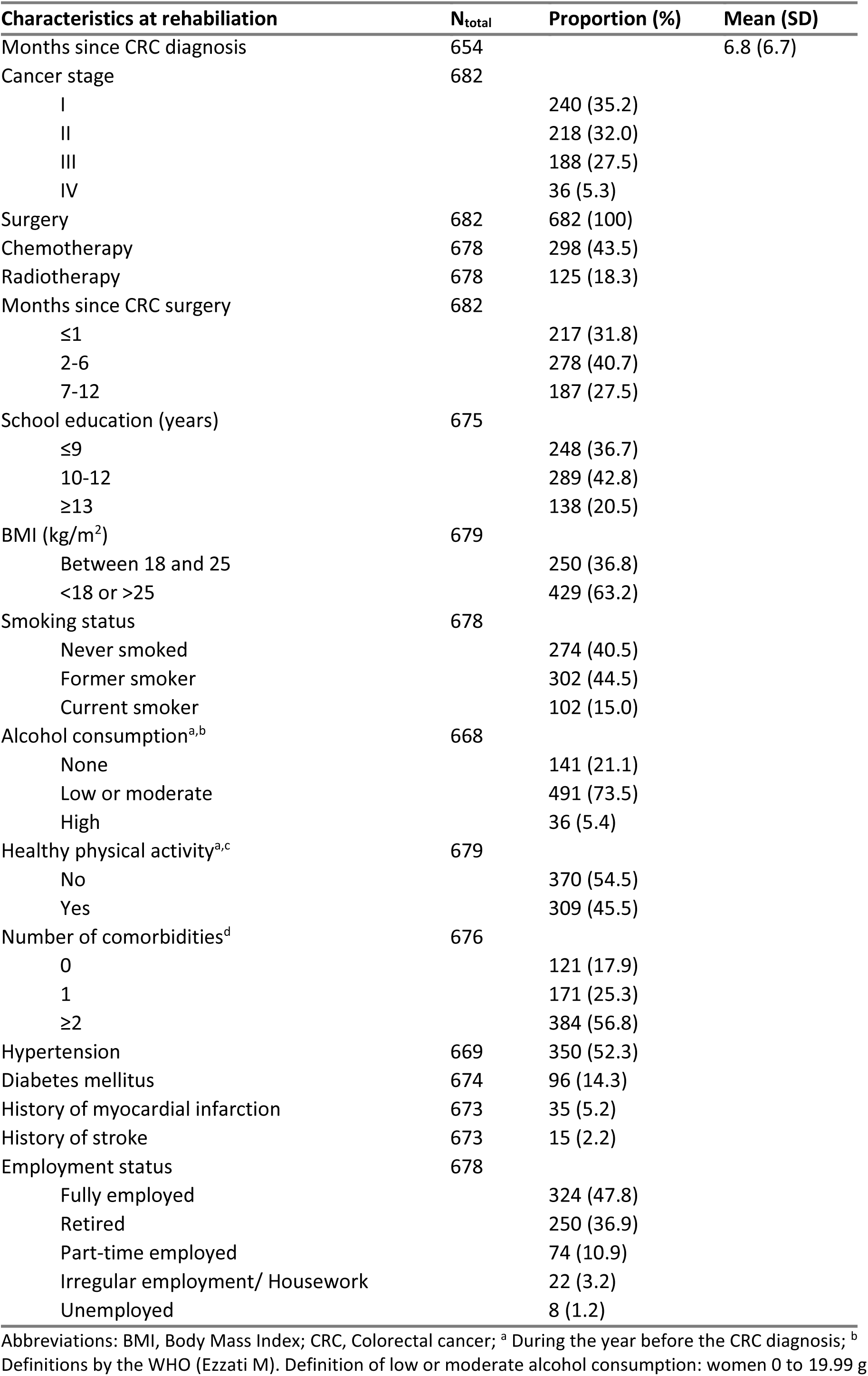
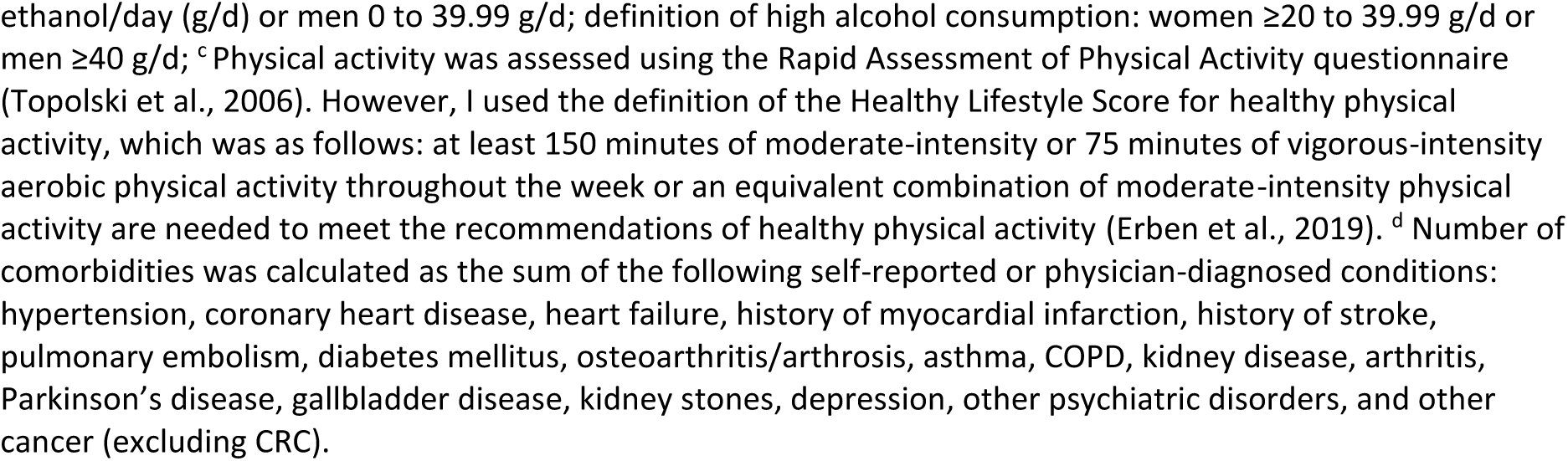
Characteristics of study participants at recruitment in German rehabilitation clinics (n=682).

Regarding lifestyle factors, 36.8% fulfilled the definition of a healthy body weight, 40.5% were non-smokers, 73.5% reported low or moderate alcohol intake, and 45.5% fulfilled the criteria for healthy physical activity. Hypertension and diabetes were present in 52.5 % and 14.3 %, respectively. In addition, 5.2% and 2.2% had a history of myocardial infarction or stroke, respectively.

Regarding employment prior to CRC diagnosis, 42.8% of participants were fully employed, 9.5% were part-time employed, 2.8% were engaged in irregular work or housework, 1.2% were unemployed, and 34.5% were retired.

### 3.2 Trajectories of the Healthy Lifestyle Score and its components

The HLS (mean [95% CI]) decreased slightly from before CRC diagnosis (2.7 [2.6–2.7]) to rehabilitation (2.6 [2.5–2.6]), and then increased to its highest level 12 months after rehabilitation (2.8 [2.7–2.9]). However, the confidence intervals overlapped, which indicates a rather stable HLS over time in CRC patients. **Figure 1** shows the distribution of the HLS at these time points, and the majority always had an intermediate HLS (2-3 points). The proportion of subjects with low HLS (0-1 point) remained similar. The only relevant change is that subjects with a high HLS (4-5 points) first decrease from before CRC diagnosis to rehabilitation (from 22% to 17%) and afterwards their proportion increases again until the 12-month-follow-up (from 17% to 29%).

**Figure 1.**
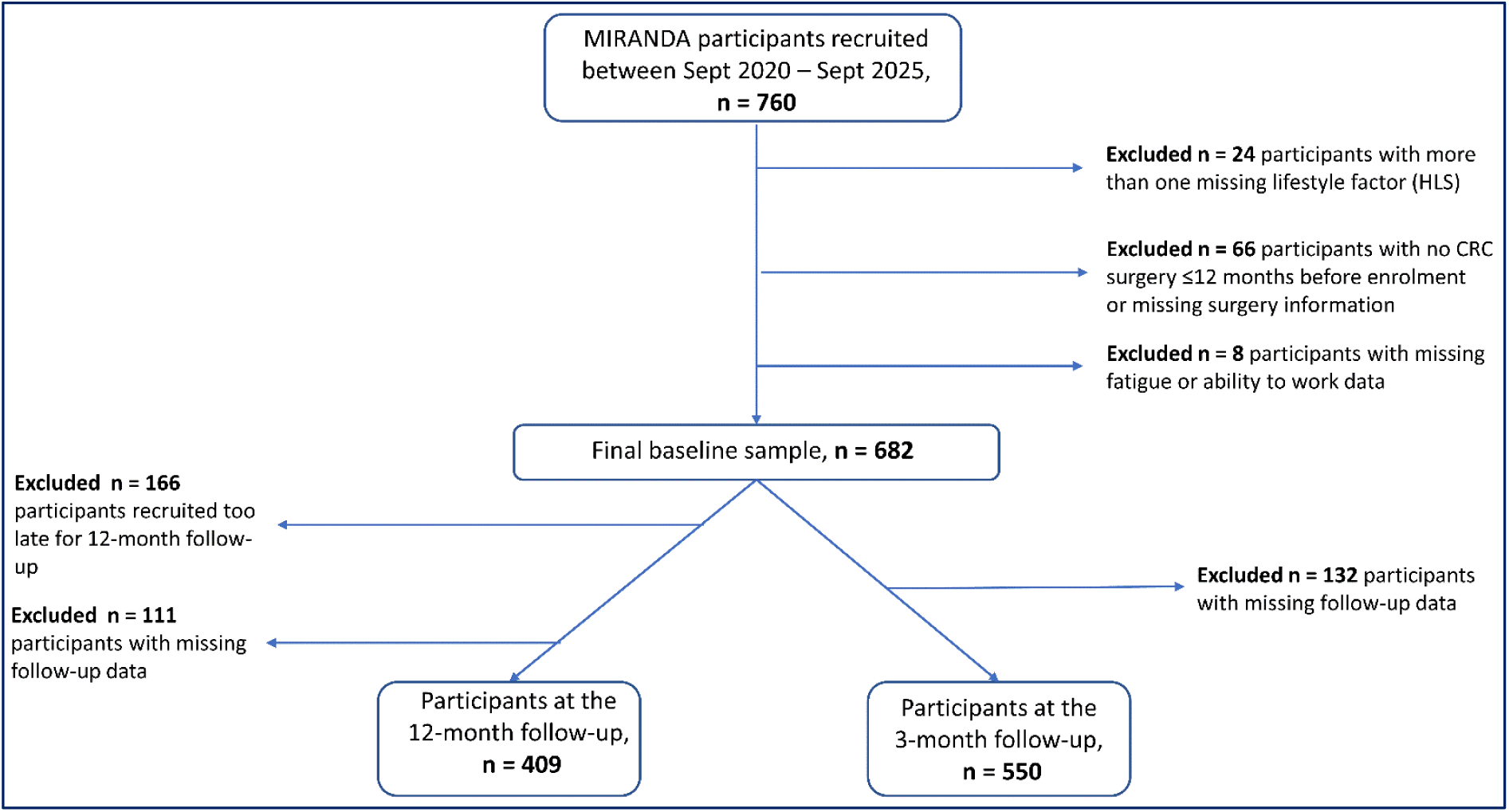
Flow chart of study participants.

**Figure 1.**
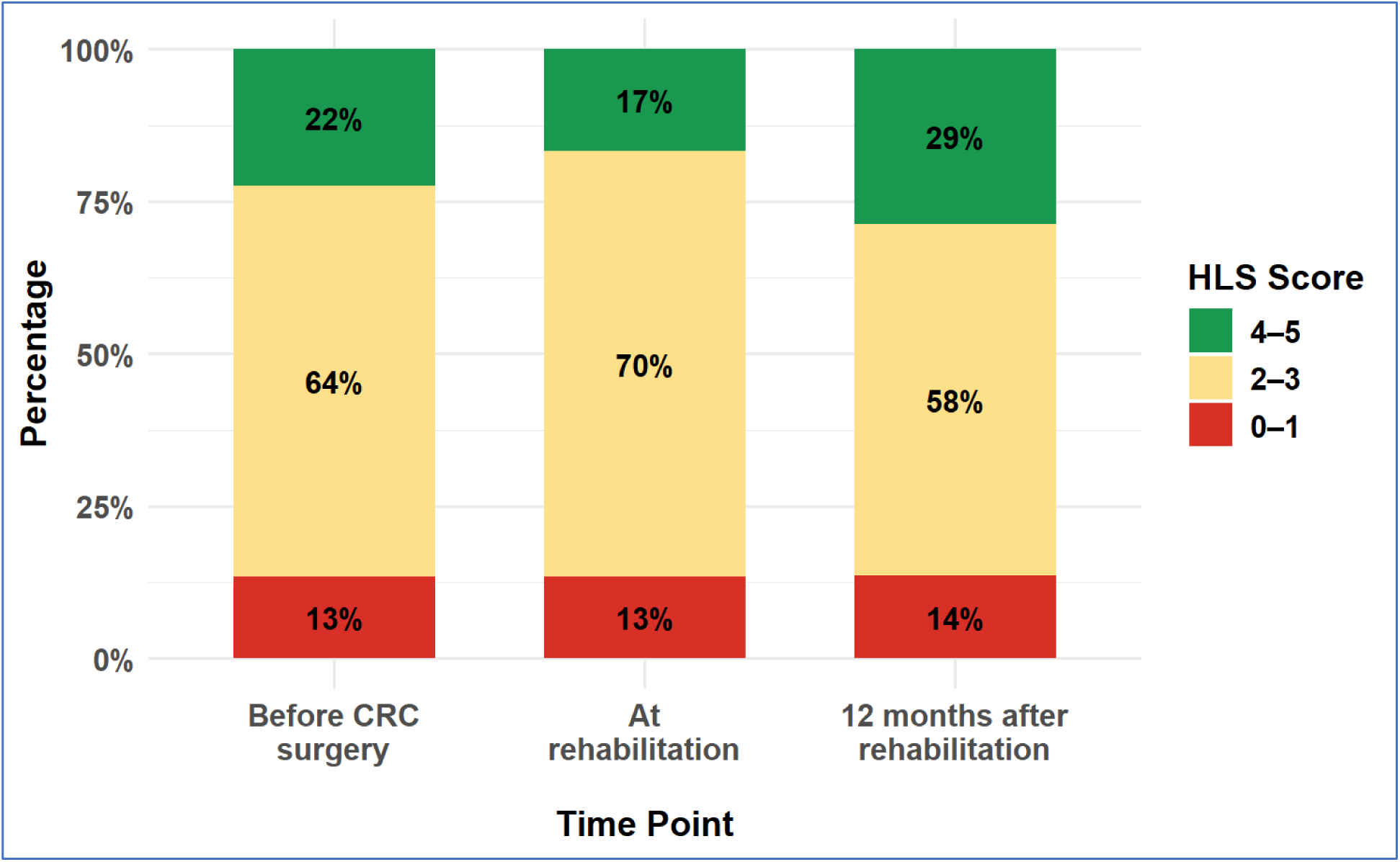
Distribution of the Healthy Lifestyle Score (HLS) before and after colorectal cancer (CRC) treatment.

The trajectories of the individual components of the HLS are shown in **Figure 2**. Most components remained relatively stable over time. Physical activity compliance decreased markedly from before diagnosis to rehabilitation and almost completely got back to its original amount twelve months after rehabilitation. The criterion of a healthy BMI had an opposite trend; it was fulfilled by more participants during rehabilitation than before CRC diagnosis and 12 months after rehabilitation. Compliance with dietary recommendations increased slightly after rehabilitation but the difference was not statistically significant because confidence intervals overlapped.

**Figure 2.**
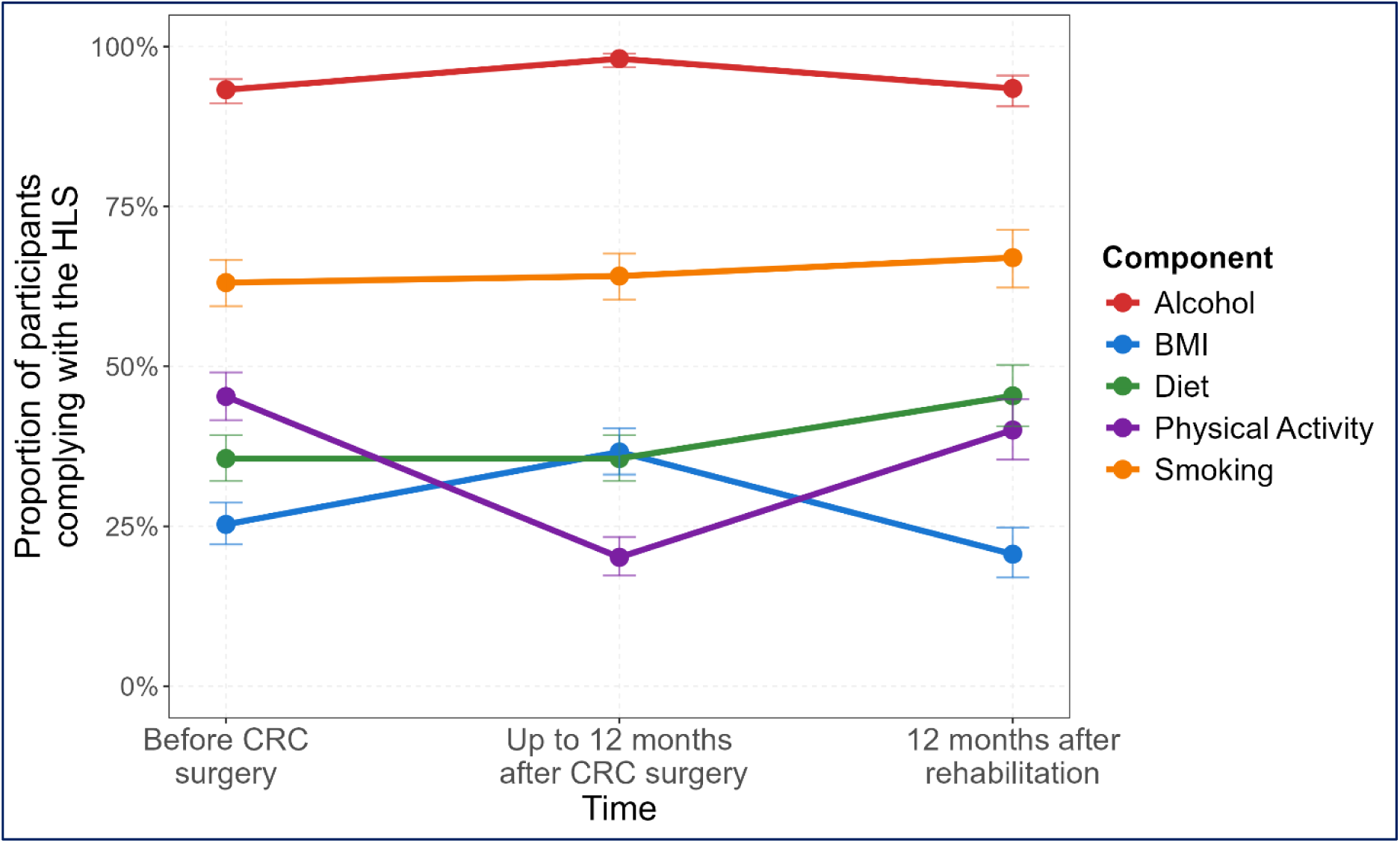
Trajectories of individual lifestyle components of the Healthy Lifestyle Score (HLS) over time.

### 3.2 Trajectories of fatigue and ability to work scores in the first year after rehabilitation

The FACIT-F-FS and FACIT-F-FWB-AW increased significantly from rehabilitation to 3 month-follow-up and plateaued thereafter (**Figure 3A and 3B**). The proportion of subjects with clinical fatigue decreased from 50.3% to 30.2% until the follow-up questionnaire 3 months after rehabilitation and was similar 12 months after rehabilitation (30.6%) (**Figure 4**). In concordance, the proportion of subjects with low-to-moderate ability to work decreased from 39.7% at rehabilitation to 24.7% at 3 months and was 18.3% at 12 months after rehabilitation (**Figure 4**).

**Figure 3.**
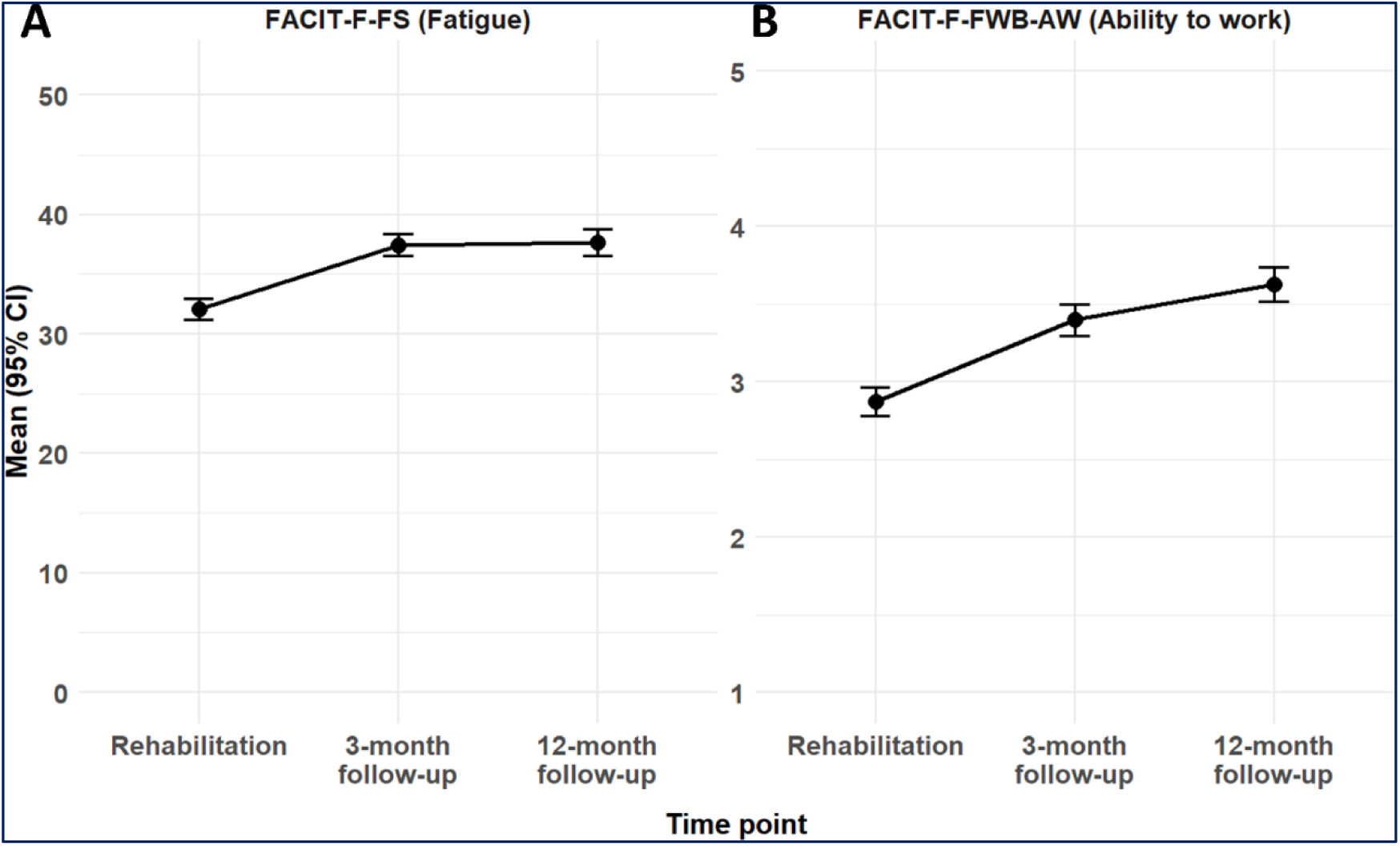
Trajectories of fatigue and ability to work during the first year after rehabilitation. (A) Mean FACIT-F-FS scores and (B) mean FACIT-F-FWB-AW scores from rehabilitation to 3-and 12-month follow-up (95% CI).

**Figure 4.**
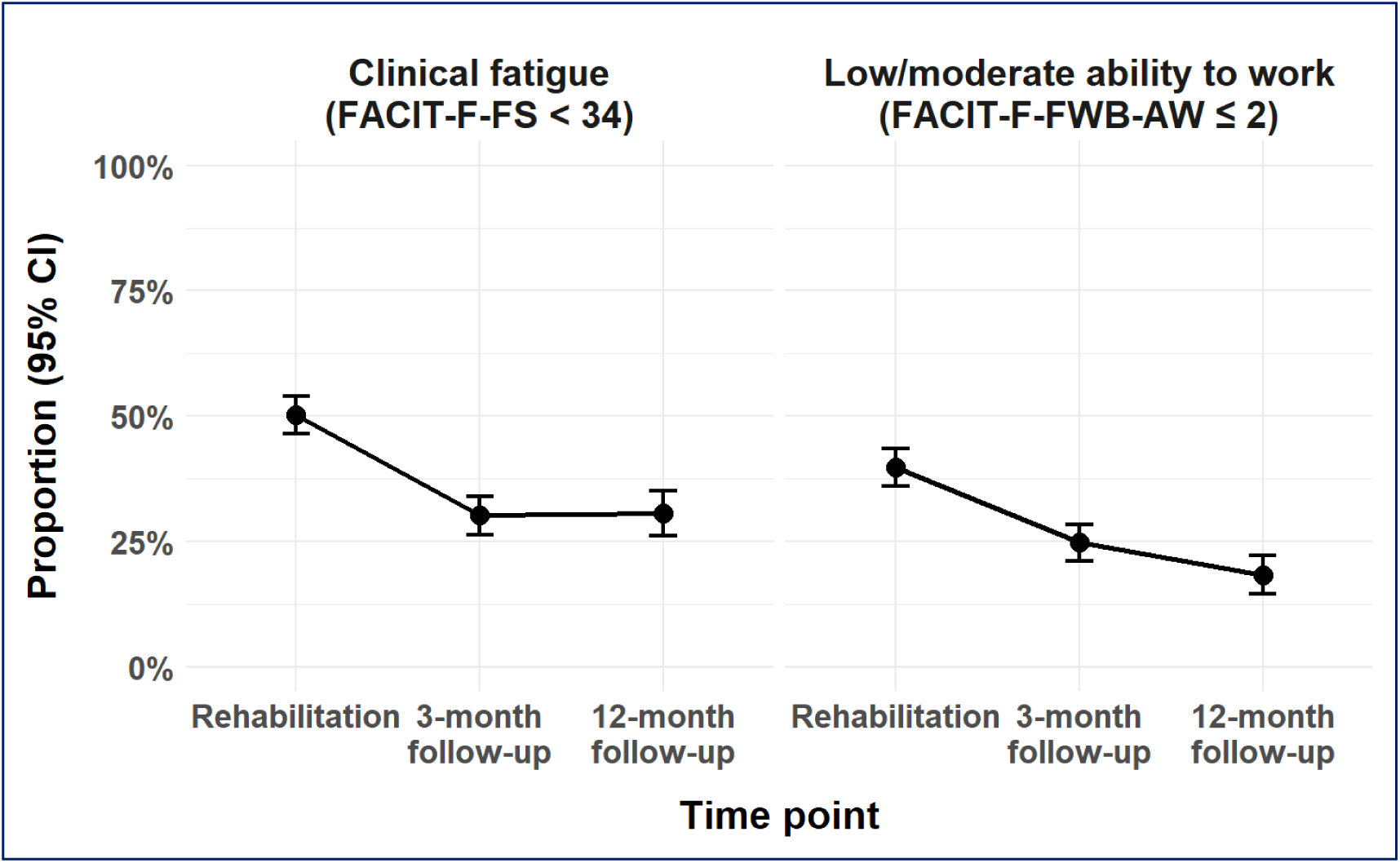
Proportion of patients with clinical fatigue (FACIT-F-FS <34) and low-to-moderate ability to work (FACIT-F-FWB-AW ≤2) over time.

### 3.3 Associations of lifestyle factors prior CRC diagnosis with cancer-related fatigue and the ability to work at rehabilitation

The associations of the pre-diagnosis HLS and individual lifestyle factors with fatigue and ability to work at rehabilitation are shown in **Table 3**. Each one-point increase in pre-diagnosis HLS was statistically significantly associated with higher FACIT-F-FS scores, indicating lower fatigue (β = 0.98, P=0.025). In addition, participants with 4–5 score points had significantly higher FACIT-F-FS scores (indicating lower fatigue), compared with those with an HLS of 0–1, (β = 4.57, P=0.004). The pre-diagnosis HLS was not significantly associated with the ability to work at rehabilitation.

**Table 3.**
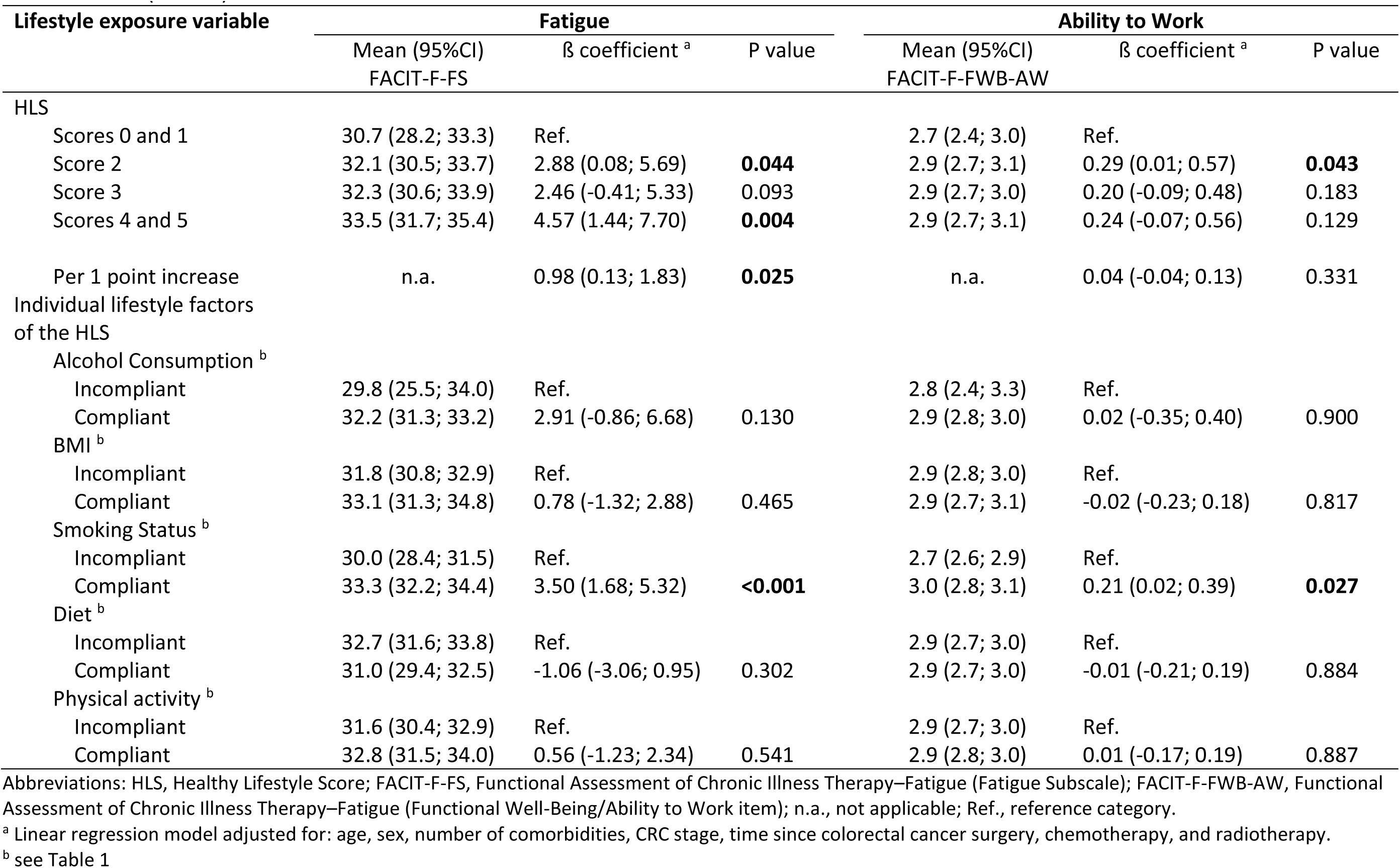
Associations of the Healthy Lifestyle Score and its single components before CRC diagnosis with fatigue and the ability to work at rehabilitation (N=682)

| Lifestyle exposure variable | Fatigue |  |  | Ability to Work |  |  |
| --- | --- | --- | --- | --- | --- | --- |
| | Mean (95%CI)<br>FACIT-F-FS | $\beta$ coefficient <sup>a</sup> | P value | Mean (95%CI)<br>FACIT-F-FWB-AW | $\beta$ coefficient <sup>a</sup> | P value |
| HLS |  |  |  |  |  |  |
| Scores 0 and 1 | 30.7 (28.2; 33.3) | Ref. |  | 2.7 (2.4; 3.0) | Ref. |  |
| Score 2 | 32.1 (30.5; 33.7) | 2.88 (0.08; 5.69) | **0.044** | 2.9 (2.7; 3.1) | 0.29 (0.01; 0.57) | **0.043** |
| Score 3 | 32.3 (30.6; 33.9) | 2.46 (-0.41; 5.33) | 0.093 | 2.9 (2.7; 3.0) | 0.20 (-0.09; 0.48) | 0.183 |
| Scores 4 and 5 | 33.5 (31.7; 35.4) | 4.57 (1.44; 7.70) | **0.004** | 2.9 (2.7; 3.1) | 0.24 (-0.07; 0.56) | 0.129 |
| Per 1 point increase | n.a. | 0.98 (0.13; 1.83) | **0.025** | n.a. | 0.04 (-0.04; 0.13) | 0.331 |
| Individual lifestyle factors<br>of the HLS |  |  |  |  |  |  |
| Alcohol Consumption <sup>b</sup> |  |  |  |  |  |  |
| Incompliant | 29.8 (25.5; 34.0) | Ref. |  | 2.8 (2.4; 3.3) | Ref. |  |
| Compliant | 32.2 (31.3; 33.2) | 2.91 (-0.86; 6.68) | 0.130 | 2.9 (2.8; 3.0) | 0.02 (-0.35; 0.40) | 0.900 |
| BMI <sup>b</sup> |  |  |  |  |  |  |
| Incompliant | 31.8 (30.8; 32.9) | Ref. |  | 2.9 (2.8; 3.0) | Ref. |  |
| Compliant | 33.1 (31.3; 34.8) | 0.78 (-1.32; 2.88) | 0.465 | 2.9 (2.7; 3.1) | -0.02 (-0.23; 0.18) | 0.817 |
| Smoking Status <sup>b</sup> |  |  |  |  |  |  |
| Incompliant | 30.0 (28.4; 31.5) | Ref. |  | 2.7 (2.6; 2.9) | Ref. |  |
| Compliant | 33.3 (32.2; 34.4) | 3.50 (1.68; 5.32) | **<0.001** | 3.0 (2.8; 3.1) | 0.21 (0.02; 0.39) | **0.027** |
| Diet <sup>b</sup> |  |  |  |  |  |  |
| Incompliant | 32.7 (31.6; 33.8) | Ref. |  | 2.9 (2.7; 3.0) | Ref. |  |
| Compliant | 31.0 (29.4; 32.5) | -1.06 (-3.06; 0.95) | 0.302 | 2.9 (2.7; 3.0) | -0.01 (-0.21; 0.19) | 0.884 |
| Physical activity <sup>b</sup> |  |  |  |  |  |  |
| Incompliant | 31.6 (30.4; 32.9) | Ref. |  | 2.9 (2.7; 3.0) | Ref. |  |
| Compliant | 32.8 (31.5; 34.0) | 0.56 (-1.23; 2.34) | 0.541 | 2.9 (2.8; 3.0) | 0.01 (-0.17; 0.19) | 0.887 |

Abbreviations: HLS, Healthy Lifestyle Score; FACIT-F-FS, Functional Assessment of Chronic Illness Therapy–Fatigue (Fatigue Subscale); FACIT-F-FWB-AW, Functional Assessment of Chronic Illness Therapy–Fatigue (Functional Well-Being/Ability to Work item); n.a., not applicable; Ref., reference category.
<sup>a</sup> Linear regression model adjusted for: age, sex, number of comorbidities, CRC stage, time since colorectal cancer surgery, chemotherapy, and radiotherapy.
<sup>b</sup> see Table 1

Among the individual lifestyle factors, compliance with the smoking criterion of the HLS was statistically significantly associated with higher FACIT-F-FS scores, indicating lower fatigue symptoms (β = 3.50, p < 0.001) and higher FACIT-F-FWB-AW scores, indicating a higher ability to work (β = 0.21, P = 0.027). The other life-style factors were not statistically significantly associated with the two outcomes.

### 3.4 Cross-sectional associations of lifestyle factors with fatigue and ability to work - all assessed during rehabilitation

The cross-sectional associations of the HLS and individual lifestyle factors with baseline fatigue and ability to work are presented in **Table 4**. Each 1-point increase in the HLS was statistically significantly associated with higher FACIT-F-FS scores (β = 1.65, p < 0.001) and higher FACIT-F-FWB-AW scores (β = 0.13, P = 0.008), indicating lower fatigue and higher ability to work. The association of compliance with the smoking criterion prior to diagnosis with fatigue and ability to work was also observed in the cross-sectional analysis. In contrast to pre-diagnosis physical activity, regular physical activity during rehabilitation was statistically significantly associated with lower fatigue (FACIT-F-FS: β = 3.89, p < 0.001) and higher ability to work (FACIT-F-FWB-AW: β = 0.28, P = 0.013), indicating lower fatigue and better ability to work. Alcohol consumption, BMI, and diet were not associated with the outcomes.

**Table 4.** Cross-sectional associations of lifestyle factors with fatigue and the ability to work at rehabilitation (N=682).

| Lifestyle exposure variable | Fatigue |  |  | Ability to Work |  |  |
| --- | --- | --- | --- | --- | --- | --- |
| | Mean (95%CI) FACIT-F-FS | $\beta$ coefficient <sup>a</sup> | P value | Mean (95%CI) FACIT-F-FWB-AW | $\beta$ coefficient <sup>a</sup> | P value |
| HLS |  |  |  |  |  |  |
| Scores 0 and 1 | 29.6 (27.4; 31.8) | Ref. |  | 2.6 (2.4; 2.8) | Ref. |  |
| Score 2 | 31.8 (30.2; 33.3) | 2.15 (-0.67; 4.96) | 0.135 | 2.8 (2.6; 2.9) | 0.14 (-0.14; 0.43) | 0.323 |
| Score 3 | 32.3 (30.7; 33.9) | 3.26 (0.33; 6.19) | **0.029** | 2.9 (2.8; 3.1) | 0.31 (0.02; 0.61) | **0.035** |
| Scores 4 and 5 | 35.0 (33.0; 37.0) | 6.14 (2.83; 9.44) | **0.001** | 3.1 (2.9; 3.3) | 0.40 (0.06; 0.73) | **0.019** |
| Per 1 point increase | n.a. | 1.65 (0.73; 2.57) | **<0.001** | n.a. | 0.13 (0.03; 0.22) | **0.008** |
| Individual lifestyle factors of the HLS |  |  |  |  |  |  |
| Alcohol Consumption <sup>b</sup> |  |  |  |  |  |  |
| Incompliant | 31.2 (22.0; 40.3) | Ref. |  | 3.0 (2.1; 3.9) | Ref. |  |
| Compliant | 32.1 (31.2; 33.0) | 1.68 (-4.95; 8.31) | 0.619 | 2.9 (2.8; 3.0) | -0.07 (-0.74; 0.59) | 0.827 |
| BMI <sup>b</sup> |  |  |  |  |  |  |
| Incompliant | 32.1 (31.0; 33.2) | Ref. |  | 2.9 (2.7; 3.0) | Ref. |  |
| Compliant | 32.2 (30.6; 33.7) | 0.25 (-1.63; 2.14) | 0.792 | 2.9 (2.8; 3.1) | 0.04 (-0.15; 0.23) | 0.656 |
| Smoking Status <sup>b</sup> |  |  |  |  |  |  |
| Incompliant | 30.0 (28.4; 31.5) | Ref. |  | 2.7 (2.6; 2.9) | Ref. |  |
| Compliant | 33.3 (32.2; 34.4) | 3.52 (1.69; 5.35) | **<0.001** | 2.9 (2.8; 3.1) | 0.19 (0.00; 0.37) | **0.045** |
| Diet <sup>b</sup> |  |  |  |  |  |  |
| Incompliant | 32.3 (31.2; 33.4) | Ref. |  | 2.8 (2.7; 2.9) | Ref. |  |
| Compliant | 31.6 (30.2; 33.1) | -0.15 (-2.07; 1.77) | 0.879 | 2.9 (2.8; 3.1) | 0.08 (-0.11; 0.27) | 0.414 |
| Physical activity <sup>b</sup> |  |  |  |  |  |  |
| Incompliant | 31.1 (30.1; 32.2) | Ref. |  | 2.8 (2.7; 2.9) | Ref. |  |
| Compliant | 35.9 (34.1; 37.8) | 3.89 (1.70; 6.08) | **<0.001** | 3.2 (3.0; 3.4) | 0.28 (0.06; 0.50) | **0.013** |

Bold print: Statistically significant (p<0.05)
Abbreviations: HLS, Healthy Lifestyle Score; FACIT-F-FS, Functional Assessment of Chronic Illness Therapy–Fatigue (Fatigue Subscale); FACIT-F-FWB-AW, Functional Assessment of Chronic Illness Therapy–Fatigue (Functional Well-Being/Ability to Work item); n.a., not applicable; Ref., reference category.
<sup>a</sup> Linear regression model adjusted for: age, sex, number of comorbidities, CRC stage, time since colorectal cancer surgery, chemotherapy, and radiotherapy.
<sup>b</sup> see Table 1

### 3.4 Longitudinal associations of pre-diagnosis lifestyle factors with cancer-related fatigue and the ability to work 3 and 12 months after rehabilitation

As the physical activity levels and the BMI are only temporarily low at rehabilitation following the CRC surgery and increase again in the first year after rehabilitation to their original states prior CRC diagnosis, we focus on the reported life-style factors prior CRC diagnosis in the longitudinal analysis. **Table 5** shows the associations of the pre-diagnosis HLS and its components with changes in FACIT-F-FS and FACIT-F-FWB-AW scores from rehabilitation to 3 months after rehabilitation in a linear regression model adjusted for the distribution of the respective score at rehabilitation and some other covariates. The total HLS was neither associated with changes in fatigue nor ability to work scores in the longitudinal analysis. Among the individual lifestyle factors, adherence to alcohol consumption recommendations before CRC diagnosis was the only factor associated with increasing FACIT-F-FS (β = 4.04, P = 0.025) and FACIT-F-FWB-AW scores (β = 0.58, P = 0.008) from rehabilitation to 3-month follow-up, indicating improved fatigue and ability to work. Compliance with other life-style factors (BMI, smoking status, diet, and physical activity) pre-diagnosis was not associated with changes in the two outcomes after rehabilitation.

**Table 5.** Longitudinal association of pre-diagnosis lifestyle factors with changes in fatigue and the ability to work from rehabilitation to 3 months after rehabilitation (N=550).

| Lifestyle exposure variable | Fatigue |  |  | Ability to Work |  |  |
| --- | --- | --- | --- | --- | --- | --- |
| | Mean change in FACIT-F-FS (95% CI) | $\beta$ coefficient <sup>a</sup> | P value | Mean change in FACIT-F-FWB-AW (95%CI) | $\beta$ coefficient <sup>a</sup> | P value |
| HLS |  |  |  |  |  |  |
| Scores 0 and 1 | 4.4 (1.7; 7.0) | Ref. |  | 0.5 (0.2; 0.8) | Ref. |  |
| Score 2 | 5.7 (4.1; 7.2) | 0.68 (-2.04; 3.41) | 0.622 | 0.5 (0.3; 0.7) | 0.00 (-0.33; 0.33) | 0.998 |
| Score 3 | 5.2 (3.8; 6.7) | 0.55 (-2.21; 3.32) | 0.695 | 0.7 (0.5; 0.8) | 0.19 (-0.14; 0.53) | 0.255 |
| Scores 4 and 5 | 5.7 (4.0; 7.3) | 0.75 (-2.27; 3.77) | 0.626 | 0.6 (0.4; 0.8) | 0.13 (-0.23; 0.50) | 0.480 |
| Per 1 point increase | n.a. | 0.34 (-0.46; 1.14) | 0.405 | n.a. | 0.06 (-0.03; 0.16) | 0.211 |
| Individual lifestyle factors of the HLS |  |  |  |  |  |  |
| Alcohol Consumption <sup>b</sup> |  |  |  |  |  |  |
| Incompliant | 1.1 (-2.7; 4.9) | Ref. |  | 0.0 (-0.5; 0.5) | Ref. |  |
| Compliant | 5.6 (4.7; 6.4) | 4.04 (0.51; 7.57) | **0.025** | 0.6 (0.5; 0.7) | 0.58 (0.15; 1.00) | **0.008** |
| BMI <sup>b</sup> |  |  |  |  |  |  |
| Incompliant | 5.4 (4.4; 6.3) | Ref. |  | 0.5 (0.4; 0.6) | Ref. |  |
| Compliant | 5.0 (3.5; 6.6) | -0.19 (-2.16; 1.78) | 0.847 | 0.6 (0.4; 0.8) | 0.05 (-0.19; 0.29) | 0.692 |
| Smoking Status <sup>b</sup> |  |  |  |  |  |  |
| Incompliant | 5.4 (4.0; 6.9) | Ref. |  | 0.5 (0.3; 0.6) | Ref. |  |
| Compliant | 5.2 (4.2; 6.2) | -0.30 (-2.02; 1.42) | 0.730 | 0.6 (0.5; 0.7) | 0.16 (-0.05; 0.37) | 0.125 |
| Diet <sup>b</sup> |  |  |  |  |  |  |
| Incompliant | 4.8 (3.8; 5.9) | Ref. |  | 0.6 (0.4; 0.7) | Ref. |  |
| Compliant | 6.2 (4.8; 7.5) | 0.67 (-1.17; 2.51) | 0.477 | 0.5 (0.4; 0.7) | -0.05 (-0.27; 0.17) | 0.638 |
| Physical activity <sup>b</sup> |  |  |  |  |  |  |
| Incompliant | 5.3 (4.2; 6.5) | Ref. |  | 0.6 (0.4; 0.7) | Ref. |  |
| Compliant | 5.3 (4.1; 6.5) | 0.45 (-1.20; 2.10) | 0.594 | 0.5 (0.4; 0.7) | -0.00 (-0.20; 0.20) | 0.973 |

Bold print: Statistically significant (p<0.05)
Abbreviations: HLS, Healthy Lifestyle Score; FACIT-F-FS, Functional Assessment of Chronic Illness Therapy–Fatigue (Fatigue Subscale); FACIT-F-FWB-AW, Functional Assessment of Chronic Illness Therapy–Fatigue (Functional Well-Being/Ability to Work item); n.a., not applicable; Ref., reference category.
<sup>a</sup> Linear regression model adjusted for: age, sex, number of comorbidities, CRC stage, time since colorectal cancer surgery, chemotherapy, radiotherapy, baseline FACIT-F-FS, and baseline FACIT-FWB-ATW.
<sup>b</sup> see Table 1

Results with fatigue and ability to work data from the assessment 12 months after rehabilitation were very similar (**Supplementary Table 1**), which can be explained by strong correlations between the 3- and 12-month assessments for both outcomes. The FACIT-F-FS showed a high Pearson correlation between 3 and 12 months (r = 0.76, 95% CI 0.72–0.80, p < 0.001), and the FACIT-F-FWB-AW showed a moderate Spearman correlation between the two time-points (r = 0.58, p < 0.001).

## 6. Discussion

### 4.1 Results summary

This study examined trajectories of healthy lifestyle indicators and their associations with cancer-related fatigue and the ability to work in CRC survivors. Overall, the Healthy Lifestyle Score (HLS) remained largely stable from before diagnosis to one year after rehabilitation, with only physical activity showing a marked temporary decline after surgery and partial recovery thereafter.

In retrospective and cross-sectional analyses, higher HLS values were consistently associated with lower fatigue and better ability to work at rehabilitation. Participants with higher HLS reported notably lower fatigue levels than those with low HLS, and each one-point increase in HLS corresponded to less fatigue and better self-rated work ability. Among the individual lifestyle factors, non-smoking and regular physical activity during rehabilitation were associated with lower fatigue, while non-smoking was also associated with higher work ability.

In longitudinal models, however, neither the pre-diagnosis nor the rehabilitation HLS predicted changes in fatigue or work ability at 3- or 12-month follow-up. Across follow-ups, adherence to alcohol-moderation recommendations prior to diagnosis was the only lifestyle component associated with improvements in both fatigue and work ability. Other lifestyle factors showed no consistent longitudinal associations.

The diagnosis of CRC often prompts significant changes in health behaviors among patients. A notable shift in health behavior among CRC patients is evident in smoking habits, with a study revealing that 62.4% of patients ceased smoking post-diagnosis [14]. Additionally, dietary changes are among the most common lifestyle adjustments made after diagnosis, while patients often express a strong desire for increased physical activity [15]. Despite this interest, a decline in physical activity levels from pre-diagnosis to post-diagnosis has been documented, underscoring the necessity for more targeted and effective interventions [16]. Furthermore, adhering to these lifestyle changes in the long term presents its own challenges. Previous study indicated that approximately 50% of long-term CRC survivors do not report significant lifestyle alterations five years post-diagnosis [11]. This highlights the challenges in maintaining lifestyle modifications over an extended period among the CRC survivor population.

### 4.2 Changes in individual lifestyle factors from before CRC diagnosis to after rehabilitation

In this cohort, engagement in regular physical activity declined markedly after surgery and partially recovered following rehabilitation. This trajectory likely reflects treatment-related reductions in mobility and endurance, consistent with previous reports showing that more than half of CRC survivors have not regained preoperative physical function six months after diagnosis [17]. The ability to sustain moderate-to-vigorous physical activity tends to recover slowly, with only modest long-term gains, unless structured interventions are implemented [18, 19]. This pattern underscores that the inpatient rehabilitation phase is a critical period for attempting to restore and improve physical function among CRC survivors.

Smoking abstinence remained stable throughout the observation period, indicating little spontaneous change in smoking behavior after diagnosis or treatment. These findings parallel prior studies showing that most cancer survivors maintain their pre-diagnosis smoking habits [20] and support the integration of targeted cessation counseling within rehabilitation programs.

Adherence to alcohol-moderation recommendations was consistently high, with only minor fluctuations across timepoints. Similar stability has been observed in other CRC cohorts, where alcohol consumption typically declines shortly after diagnosis and then plateaus [21–23]. Although earlier guidelines permitted limited consumption, current World Cancer Research Fund and American Institute for Cancer Research (WCRF/AICR) recommendations advise complete abstinence [24].

Dietary adherence remained relatively unchanged from before diagnosis to one year after rehabilitation, underscoring the persistence of dietary habits. Previous research also shows that only a minority of CRC survivors make lasting dietary adjustments [25, 26]. Even when short-term improvements are achieved through structured interventions, these gains often diminish within two years [27, 28]. Sustained improvements, therefore, appear to require continuous, personalized nutritional counseling.

### 4.3 Interpretation of the HLS (HLS score) findings

Higher pre-diagnosis and post-surgery HLS values were associated with lower fatigue and better ability to work at rehabilitation. Participants meeting more healthy-behavior criteria reported markedly lower fatigue levels than those with low HLS, and each one-point increase in HLS corresponded to gradual improvements in fatigue and work ability. These findings align with evidence from large CRC cohorts showing that composite lifestyle indices are strongly related to health-related quality of life, including fatigue subscales [11], and with meta-analytic data across cancer types indicating that integrated lifestyle scores correlate more closely with psychosocial outcomes than single behaviors [29]. Together, these results underscore that cumulative adherence to multiple health behaviors, rather than isolated habits, best captures the relationship between lifestyle and fatigue.

In longitudinal analyses, HLS did not predict fatigue or ability to work at 3- or 12-month follow-up, suggesting that the impact of lifestyle behaviors before or during treatment is more pronounced on a patient’s symptom burden at that time rather than on their short-term recovery trajectory.

### 4.4 Individual lifestyle factors and fatigue

#### 4.4.1 Smoking status

CRC patients who never smoked or had less than 30 pack-years before diagnosis reported lower fatigue than those who continued to smoke. Similar associations have been observed across large survivor cohorts, where current smokers exhibit greater overall symptom burden, including fatigue [30]. Smoking contributes to oxidative stress, systemic inflammation, and sleep disruption—mechanisms central to cancer-related fatigue [31]. Conversely, cessation improves pulmonary function and reduces inflammation [32]. Meta-analytic data further confirm smoking as a modifiable determinant of fatigue [33], reinforcing the importance of integrating structured cessation programs into CRC rehabilitation.

#### 4.4.2 Physical activity

Participants who maintained regular physical activity during rehabilitation reported substantially less fatigue than their inactive peers, consistent with prospective evidence that greater leisure-time and moderate-to-vigorous activity before and after diagnosis predicts reduced fatigue for up to a decade post-treatment [16]. Exercise mitigates systemic inflammation, preserves muscle mass, and prevents deconditioning—key pathways underlying fatigue [34]. Randomized trials of resistance and aerobic training likewise demonstrate significant reductions in fatigue and improved quality of life during adjuvant therapy [35]. These data highlight the role of structured exercise as an essential component of CRC rehabilitation.

#### 4.4.3 Alcohol intake, diet, and body mass index

Adherence to alcohol-moderation recommendations prior to CRC diagnosis was the only lifestyle factor associated with improvements in both fatigue and work ability during follow-up. Although evidence linking alcohol to fatigue remains limited, heavy alcohol intake is known to induce systemic inflammation through Toll-like receptor 4 activation in the liver and brain [34], mechanisms also implicated in fatigue pathophysiology. Evidence on alcohol and fatigue remains limited, although heavy alcohol consumption can induce systemic inflammation through Toll-like receptor 4 activation in the liver and brain [36], mechanisms also implicated in fatigue pathophysiology [4]. Thus, our findings may reflect a non-linear relationship, where moderate consumption—potentially as a proxy for social engagement or overall wellbeing—correlates with lower fatigue, while higher intake may confer metabolic and inflammatory risks.

Dietary inflammatory load and anti-inflammatory dietary interventions have been associated with fatigue severity in other cancer populations [34, 37], but such effects were not observed in our cohort. This may reflect limitations in our dietary assessment approach, particularly the dichotomized scoring of dietary habits, which may lack the granularity needed to capture dose–response patterns or qualitative differences in nutritional intake.

Similarly, BMI may inadequately capture body composition–related mechanisms influencing fatigue: higher subcutaneous adiposity and lower muscle radiodensity have been linked to greater fatigue at diagnosis, while improvements in muscle function predict reduced fatigue during recovery [38, 39]. Future studies using objective, longitudinal measures such as accelerometry and advanced imaging are needed to delineate these relationships better.

### 4.5 Implications for rehabilitation and survivorship care

The findings of this study highlight several actionable targets for rehabilitation and long-term survivorship management. The strong associations of physical activity and non-smoking with lower fatigue underscore the importance of integrating structured exercise programs and smoking-cessation support into standard CRC rehabilitation. Exercise prescriptions should extend beyond the inpatient setting, with tailored follow-up and remote supervision to help survivors maintain gains in physical function and fatigue reduction.

Although composite HLS values were not predictive of fatigue improvement at follow-up, the stability of lifestyle patterns across time suggests that early intervention—ideally before or immediately after treatment—may be critical. Embedding brief lifestyle assessments within oncologic care could help identify individuals at risk for persistent fatigue and guide timely referral to multidisciplinary support, including physiotherapists, nutritionists, and psycho-oncologists.

The limited change in dietary adherence and the high prevalence of overweight observed in this cohort indicate that one-off counseling during rehabilitation is insufficient to achieve sustained behavioral change. Continuous, personalized lifestyle counseling supported by digital or community-based follow-up may help reinforce long-term adherence. Finally, as fatigue and work ability are interrelated yet influenced by psychosocial and behavioral factors, future rehabilitation programs should adopt a comprehensive approach combining physical activity promotion, psychological support, and vocational reintegration strategies.

### 4.6 Strengths and limitations

This study has several important strengths. It was based on a well-characterized cohort of colorectal cancer survivors with prospective follow-up and detailed information on lifestyle and patient-reported outcomes. Using a validated HLS that combined five key behaviors allowed a broad assessment of how multiple lifestyle factors together relate to cancer-related fatigue and the ability to work. Repeated follow-up assessments also made it possible to examine how changes in lifestyle over time might relate to later outcomes.

Some limitations should be considered. Fatigue and lifestyle behaviors were self-reported, which may introduce recall errors or misclassification. Lifestyle habits may also have changed after diagnosis, though pre-diagnosis data were used when available to limit this effect. The use of binary indicators for adherence may not fully capture gradual or dose-related differences. The number of participants declined at later follow-ups, reducing statistical power for longitudinal analyses. Finally, unmeasured factors such as treatment side effects, mental health, or social support could have influenced the results, even though key demographic and clinical variables were included in the models.

## 7. Conclusion

In this cohort of colorectal cancer survivors, adherence to a healthy lifestyle—particularly maintaining regular physical activity, reducing alcohol intake and abstaining from smoking—was associated with lower fatigue and better ability to work at rehabilitation baseline. These associations were evident for both pre-diagnosis and post-surgery behaviors, suggesting that sustained engagement in multiple healthy habits contributes to better functional recovery and well-being during the rehabilitation phase.

While the overall HLS remained largely stable from diagnosis to one year after rehabilitation, changes in individual components, especially the temporary decline and subsequent recovery of physical activity, highlight the impact of treatment on survivors’ capacity to maintain healthy behaviors. The absence of strong longitudinal associations indicates that short-term recovery may depend more on clinical and psychosocial factors than on lifestyle behaviors alone.

Taken together, these findings emphasize the need for early, sustained, and individualized lifestyle support as part of comprehensive colorectal cancer rehabilitation. Interventions that combine exercise programs, smoking cessation, and long-term behavioral counseling may help improve fatigue outcomes and enhance the return-to-work potential of CRC patients.

## Data Availability Statement

The data will not be published on an open-access platform. After the study is completed, interested scientists can request data use and receive pseudonymized data upon approval of this application by the principal investigator, Prof. Dr. Ben Schöttker.

## Ethics declarations

The study was conducted in accordance with the Declaration of Helsinki and in accordance with all applicable legal and regulatory requirements in Germany. The study was approved by the responsible Ethical Committee of the Faculty of Medicine Heidelberg (ethical approval code S-905/2019, date of approval: January 27 2020).

## Author Contributions

BS accounted for conceptualization, data curation, funding acquisition, project administration, reviewing and editing of the manuscript, as well as supervision of the study. TV accounted for data curation, formal analysis, methodology, and drafting the original manuscript. All co-authors accounted for the reviewing & editing of the manuscript. TV and BS are accountable for all aspects of the work in ensuring that questions related to the accuracy or integrity of any part of the work are appropriately investigated and resolved.

