## Supplemental Table 1 for "The associations of lifestyle factors with fatigue and the ability to work in the first year after colorectal cancer surgery and rehabilitation"

**Suppl. Table 1.** Associations of pre-diagnosis lifestyle factors with changes in fatigue and the ability to work from rehabilitation to 12 month after rehabilitation (N=409)

| **Lifestyle exposure variable** | **Fatigue** | | |  | **Ability to Work** | | |
| --- | --- | --- | --- | --- | --- | --- | --- |
|  | Mean (95%CI) change of FACIT-F-FS | ß coefficient ^a^ | P value |  | Mean change of FACIT-F-FWB-AW (95%CI) | ß coefficient ^a^ | P value |
| HLS |  |  |  |  |  |  |  |
| Scores 0 and 1 | 4.0 (1.1; 6.8) | Ref. |  |  | 0.6 (0.2; 1.0) | Ref. |  |
| Score 2 | 6.2 (4.2; 8.1) | 0.87 (-2.43; 4.17) | 0.606 |  | 0.8 (0.6; 1.0) | 0.17 (-0.23; 0.57) | 0.402 |
| Score 3 | 5.6 (3.9; 7.4) | 0.88 (-2.47; 4.23) | 0.606 |  | 0.9 (0.7; 1.1) | 0.26 (-0.15; 0.67) | 0.211 |
| Scores 4 and 5 | 4.6 (2.5; 6.7) | -0.55 (-4.15; 3.04) | 0.763 |  | 0.6 (0.3; 0.8) | -0.03 (-0.47; 0.41) | 0.889 |
| Per 1 point increase | n.a. | 0.03 (-0.92; 0.98) | 0.950 |  | n.a. | 0.00 (-0.12; 0.11) | 0.971 |
| Individual lifestyle factors  of the HLS |  |  |  |  |  |  |  |
| Alcohol Consumption ^b^ |  |  |  |  |  |  |  |
| Incompliant | 0.7 (-3.3; 4.7) | Ref. |  |  | 0.2 (-0.5; 0.9) | Ref. |  |
| Compliant | 5.5 (4.5; 6.5) | 4.74 (0.26; 9.23) | **0.038** |  | 0.8 (0.6; 0.9) | 0.65 (0.11; 1.20) | **0.019** |
| BMI ^b^ |  |  |  |  |  |  |  |
| Incompliant | 5.2 (4.0; 6.3) | Ref. |  |  | 0.7 (0.6; 0.9) | Ref. |  |
| Compliant | 5.7 (3.7; 7.6) | 0.59 (-1.88; 3.06) | 0.638 |  | 0.8 (0.5; 1.0) | 0.02 (-0.28; 0.32) | 0.886 |
| Smoking Status ^b^ |  |  |  |  |  |  |  |
| Incompliant | 5.6 (3.6; 7.7) | Ref. |  |  | 0.7 (0.5; 0.9) | Ref. |  |
| Compliant | 5.2 (4.1; 6.3) | -0.64 (-2.77; 1.49) | 0.558 |  | 0.7 (0.6; 0.9) | 0.04 (-0.22; 0.30) | 0.777 |
| Diet ^b^ |  |  |  |  |  |  |  |
| Incompliant | 5.4 (4.2; 6.7) | Ref. |  |  | 0.9 (0.7; 1.0) | Ref. |  |
| Compliant | 5.1 (3.4; 6.7) | -0.99 (-3.25; 1.26) | 0.388 |  | 0.5 (0.3; 0.7) | -0.29 (-0.56; -0.01) | **0.041** |
| Physical activity ^b^ |  |  |  |  |  |  |  |
| Incompliant | 5.5 (4.1; 6.9) | Ref. |  |  |  | Ref. |  |
| Compliant | 5.0 (3.6; 6.5) | 0.16 (-1.85; 2.16) | 0.878 |  |  | 0.04 (-0.20; 0.29) | 0.745 |

Bold print: Statistically significant (p<0.05)

Abbreviations: HLS, Healthy Lifestyle Score; FACIT-F-FS, Functional Assessment of Chronic Illness Therapy–Fatigue (Fatigue Subscale); FACIT-F-FWB-AW, Functional Assessment of Chronic Illness Therapy–Fatigue (Functional Well-Being/Ability to Work item); n.a., not applicable; Ref., reference category.

^a^ Linear regression model adjusted for: age, sex, number of comorbidities, CRC stage, time since colorectal cancer surgery, chemotherapy, radiotherapy, baseline FACIT-F-FS, and baseline FACIT-FWB-ATW.

^b^ see Table 1
